# Obesity and COVID-19 Mortality A Cross-Country Analysis

**DOI:** 10.1101/2021.01.28.21249723

**Authors:** Giacomo De Giorgi, Felix Michalik

## Abstract

We highlight a robust correlation between COVID-19 mortality and obesity prevalence using available country level data on COVID-19 mortality as of August 10, 2020. Such association is robust to controlling for other potential comorbidity factors: diabetes, cardiovascular, and respiratory diseases, further to a set of demographics, urban, and economic, and containment policies controls. We estimate that .6 log point increase in obesity prevalence, or 1 standard deviation, is associated with about an extra .9 log point per 100,000 deaths (or 50% of a standard deviation, .5*σ)*.

## 1 Introduction

The novel COVID-19 has so far (as of August 10, 2020) claimed more than 700,000 lives worldwide while there are about 20 million confirmed cases (Dong et al., 2020). A number of papers in the CEPR COVID series have investigated various aspects of COVID-19 impacts and mitigation strategies.^1^ However, somewhat less attention has been devoted to the role of comorbidities which could be crucial in determining the mortality risk within age groups. A relevant dimension of such comorbidities is obesity. As we know, obesity is linked to many diseases and heightened mortality risk overall (WHO, 2020). We, therefore, investigate whether obesity prevalence is linked to higher COVID-19 mortality across the world. Our cross-country analysis reveals that obesity prevalence is associated with higher COVID-19 mortality, and is so across all income quartiles of the countries for which we have data for. Such association does not disappear when conditioning on the prevalence of diabetes, cardiovascular diseases, and respiratory diseases further to share of 65 years and older, urban density, GDP per capita and containment policies. Our analysis is at the country level as we have no access to individual-level records at this stage, and we believe that such analysis provide a complementary picture to individual-level studies.

In the past few months, it has become apparent that many of the COVID-19 related deaths are in the older population with often multiple pre-existing conditions.^2^ In particular, the most cited comorbidity conditions have been identified in: i. cardiovascular diseases, ii. diabetes, iii. pre-existing respiratory conditions. In this article we investigate the relevance of an ulterior condition: obesity. Obesity is defined as a Body Mass Index (BMI) greater than or equal to 30 as per standard definition of the World Health Organization (WHO), where an individual’s BMI is weight in kilograms divided by the square of height in meters. Our analysis relates the COVID-19 deaths per 100,000 to the prevalence of obesity in the country, at the same time we account for the role of other co-factors as the ones mentioned earlier as well as other economic conditions like per capita GDP.

Some very recent work has mentioned the possibility of obesity as an important co-factor in death rates using relatively small samples, from the UK and Southern California, and has investigated the possible role of obesity as a determinant of mortality for younger patients, see Williamson et al., 2020; Tartof et al., 2020 and the literature cited therein.

Our aim here is to highlight an important dimension of heterogeneity in terms of obesity prevalence and COVID-19 mortality for all the countries we have data for (at the time of writing 138 countries). While we don’t have individual patients’ data our analysis spans all the countries with reliable data on COVID-19 cases and mortality, further to the crucial controls both in terms of health and socio-economics. A noticeable advantage of our analysis is that we cover a large number of countries spanning the entire spectrum of the world income distribution, and at the same time our analysis isn’t mediated by hospitalizations’ decisions which would inevitably define the nature of the sample of an individual level analysis (a sample selection based on observables and unobservables).

Just to give a sense of the magnitude of this association: an increase in obesity prevalence by .6 log points (or 1*σ*, Sweden vs. United Kingdom prevalences) is associated with an increase in mortality of about .9 log points of per 100,000 deaths or about 75% of the mean (about .5*σ*). This association is rather large in magnitude and translates into an extra 10 per 100,000 deaths (basically a doubling of the mean deaths per 100,000 across countries).

The fact that obesity is associated with adverse health outcomes has been documented by a large literature: for general health (e.g., GBD 2015 Obesity Collaborators, 2017; Guh et al., 2009; Flegal et al., 2013) and are major risk factors for multiple diseases, including cardiovascular diseases (e.g., Asia Pacific Cohort Studies Collaboration, 2004; Emerging Risk Factors Collaboration et al., 2011; Lu et al., 2013), type 2 diabetes (e.g., Kahn et al., 2006; Eckel et al., 2011), certain types of cancer (e.g., Renehan et al., 2008), obstructive sleep apnea (e.g., Schwartz et al., 2008), osteoarthritis (e.g., Lohmander et al., 2009; Yusuf et al., 2010), and depression (e.g., Luppino et al., 2010). It is perhaps then not surprising that obesity and COVID-19 mortality present a robust association. As mentioned earlier obesity is associated with leading causes of preventable, premature death, and recently their prevalence has been dramatically increasing, particularly in low- and middle-income countries (Abarca-GÓmez et al., 2017). The remainder of the paper proceeds as follows: Section 2 presents some of the related medical literature and a basic graphic analysis of the association between obesity prevalence at the country level; Section 3 probes the robustness of this correlation in a regression framework where several controls are added sequentially; finally, Section 4 concludes.

## 2 Association of Obesity Prevalence and Covid-19 Mortality

Recalling the impact of obesity on mortality from H1N1 influenza, there is reason to believe that obesity is one of the preexisting conditions associated with death (Dietz and Santos-Burgoa, 2020). There are multiple ways in which obesity may negatively impact COVID-19 infection by lowering cardiorespiratory as well as metabolic reserves through cardiovascular, respiratory, metabolic, and thrombotic comorbidities; and facilitating immune hyperreactivity, due to greater viral exposure and excess adipose tissue amplifying the immune response Sattar et al. (2020). While data on metabolic parameters, including BMI, in patients with COVID-19 is limited, several studies from individual cities and hospitals suggest that obesity is associated with more severe COVID-19 processes (Stefan et al., 2020). Young patients generally are at low risk of severe COVID-19 infection. Especially for this low-risk group, however, obesity might be a key risk factor. Data from an academic hospital system in New York City (see Lighter et al., 2020) and university hospitals at Johns Hopkins, University of Cincinnati, New York University, University of Washington, Florida Health, and University of Pennsylvania (see Kass et al., 2020) suggest that in populations with a high prevalence of obesity younger age groups will be more affected than in populations with a low prevalence. More recently Williamson et al., 2020; Tartof et al., 2020 use larger study population albeit limited to specific geographic areas.

Given the lack of wide-scale patient-level data with BMI records, we conduct an analysis at the country level for the 138 countries we have available and reliable data for. In Figure 1 we plot the relationship between COVID-19 mortality per 100,000 population and obesity prevalence for the 138 countries which had more than 10 COVID-19 related deaths. As one can see clearly there is a marked positive relationship, even if we exclude some influential observations such as we do in the Appendix Figure B.1. Let us be clear here, this is not a causal statement as we just present a simple association analysis here: we estimate a coefficient of 1.7 (statistically significant at the 1% level (robust t-stat: 8.3)).

**Figure 1:**
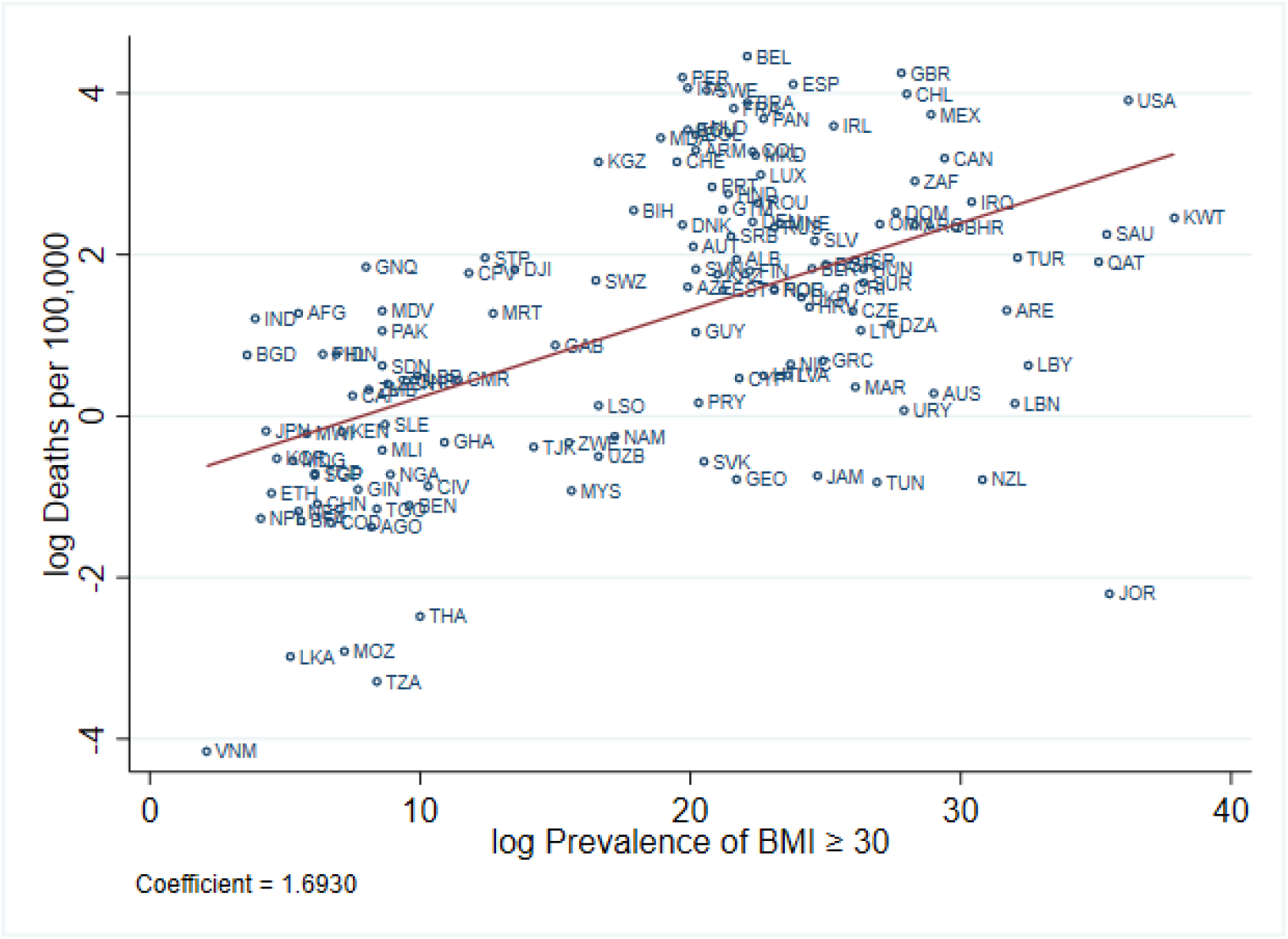
Obesity and COVID-19 Deaths per 100,000. *Notes:* Data on COVID-19 deaths from August 10, 2020.

Nevertheless, just to give a sense of the magnitude of the plotted association: an increase in obesity prevalence by 1 log point (which is, e.g., the difference between the prevalence of Indonesia and Switzerland) is associated with an increase in mortality of about 1.7 log points of per 100,000 extra deaths.

It is also quite noticeable from Figure 2 how such association is robust across income quartiles. If we think of an increase of 1 log point in obesity prevalence this is associated with 1.2, 1.4, 1.7, and 1.3 log points of extra deaths per 100,000 in the 1st, 2nd, 3rd, and 4th income quartile, respectively. Notice that those associations are not statically different from each other at conventional levels. However, we also point out how for the 4th quartile Japan, Singapur, and South Korea are influential observations as they are way low in obesity prevalence and mortality. Considering that the highest mortality rate per 100,000 at the time of writing (data up to August 10, 2020) is that of Belgium at 4.5 log points, with an EU average of about 3.1 this is a pretty large magnitude. Another way to see this is to consider moving down the prevalence rate of obesity from the US to Japan this is associated with about 2.8 log points per 100,000 fewer deaths. Once more, these are only associations with no aim at making causal statements. In the next section we will present a regression analysis where we partial out some of the possible confounding factors, i.e. other comorbidities, age structure, GDP per capita, urban density, policy responses, duration of the pandemic at the country level etc..

**Figure 2:**
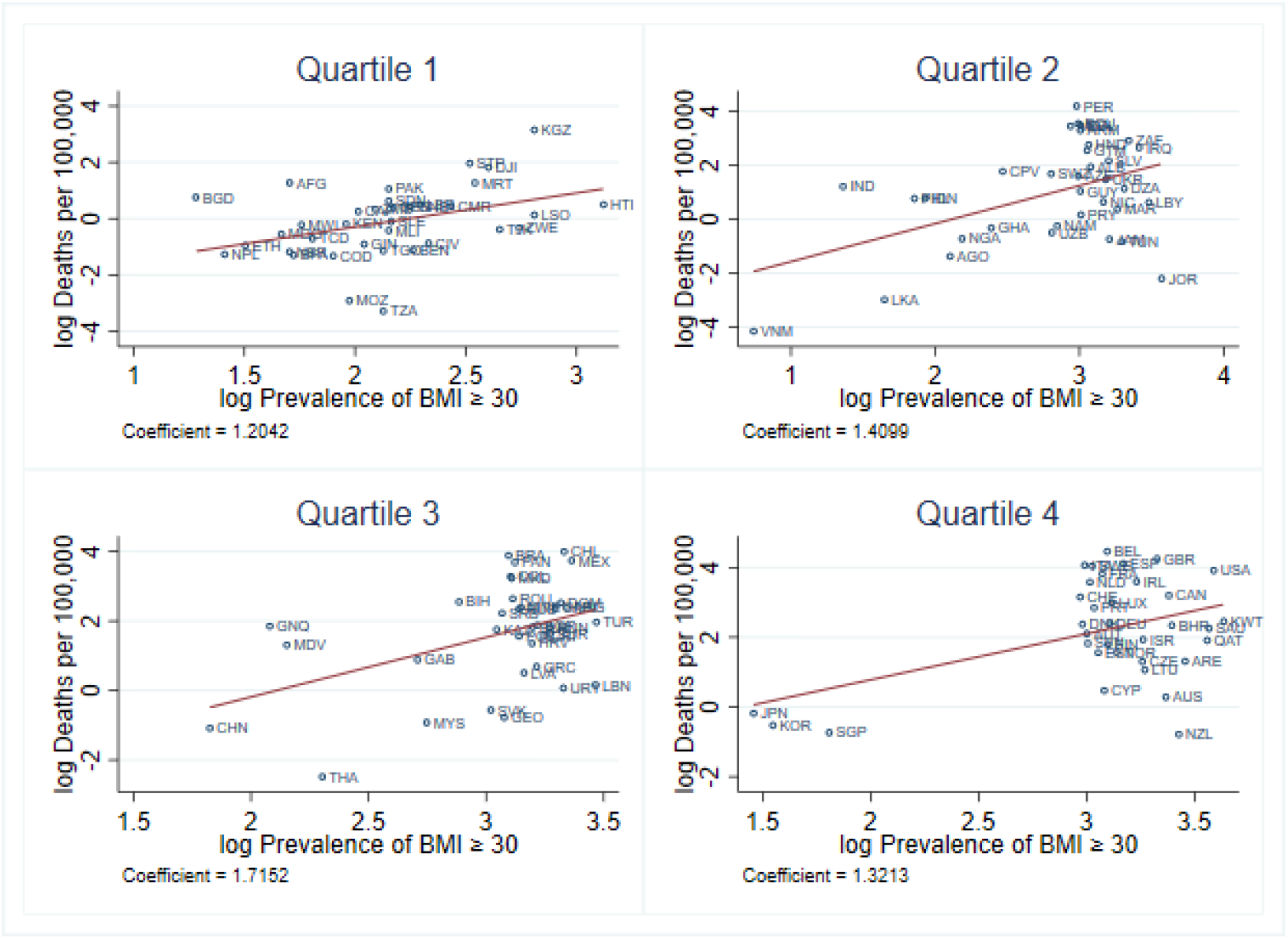
Correlation of Obesity and COVID-19 deaths, Income Quartiles. *Notes:* Data on COVID-19 deaths from August 10, 2020.

As European countries have been ahead of the diffusion and mortality curve, and somewhat took early actions, we provide in Appendix Figure C.1 a similar graphical analysis where we split the sample of countries between Europe and the Rest of the World.

## 3 Obesity and COVID-19 Mortality Controlling for Prevalence of Diabetes, Cardiovascular, and Respiratory Conditions

As discussed above, it is well known that high obesity prevalence is associated with high diabetes and cardiovascular disease prevalence. We, therefore, conduct a correlation analysis below, where we control for the prevalences of diabetes and cardiovascular diseases further to respiratory conditions at the country level, as well as controlling for the share of 65 or older in the population, GDP per capita, share of urban population, number of days since the 100th recorded case (and its square as well as cube) to control for the dynamic aspects linking infection and mortality, and timing of lockdown (see note to Table 1 and Appendix A).

**Table 1:** COVID-19 Mortality per 100,000 Population

| Date | (1)<br>Aug 10 2020 | (2)<br>Aug 10 2020 | (3)<br>Aug 10 2020 | (4)<br>Aug 10 2020 | (5)<br>Aug 10 2020 | (6)<br>Aug 10 2020 | (7)<br>Aug 10 2020 |
| --- | --- | --- | --- | --- | --- | --- | --- |
| log $BMI \geq 30$ | 1.511***<br>(0.226) | 1.494***<br>(0.303) | 1.512***<br>(0.322) | 1.508***<br>(0.265) | 1.543***<br>(0.327) | 1.486***<br>(0.341) | 1.433***<br>(0.381) |
| $Age \geq 65$ | | 0.0297<br>(0.0276) | 0.0472<br>(0.0618) | 0.0371<br>(0.0362) | 0.0503<br>(0.0602) | 0.0386<br>(0.0597) | 0.0373<br>(0.0606) |
| log Diabetes |  | -0.158<br>(0.489) |  |  | -0.0374<br>(0.552) | -0.144<br>(0.584) | -0.146<br>(0.584) |
| log Cardiovascular diseases |  |  | -0.341<br>(0.854) |  | -0.228<br>(0.948) | -0.0508<br>(0.976) | -0.0326<br>(0.979) |
| log CRDs |  |  |  | -0.356<br>(0.676) | -0.264<br>(0.853) | -0.321<br>(0.870) | -0.300<br>(0.884) |
| GDP pc / 1000 |  |  |  |  |  | 0.00750<br>(0.00745) | 0.00695<br>(0.00772) |
| log Urban population |  |  |  |  |  |  | 0.129<br>(0.455) |
| $Days > 100$ | -0.0292<br>(0.0663) | -0.0229<br>(0.0603) | -0.0178<br>(0.0598) | -0.0208<br>(0.0605) | -0.0210<br>(0.0607) | -0.0150<br>(0.0578) | -0.0167<br>(0.0583) |
| $(Days > 100)^2$ | 0.000359<br>(0.000672) | 0.000307<br>(0.000605) | 0.000273<br>(0.000598) | 0.000294<br>(0.000609) | 0.000298<br>(0.000602) | 0.000249<br>(0.000565) | 0.000265<br>(0.000569) |
| $(Days > 100)^3$ | -8.95e-07<br>(2.09e-06) | -8.27e-07<br>(1.88e-06) | -7.55e-07<br>(1.85e-06) | -8.10e-07<br>(1.89e-06) | -8.19e-07<br>(1.85e-06) | -7.44e-07<br>(1.71e-06) | -8.05e-07<br>(1.72e-06) |
| Lockdown response | 0.000186<br>(0.00210) | -0.000249<br>(0.00220) | -0.000232<br>(0.00216) | -0.000141<br>(0.00217) | -0.000216<br>(0.00222) | -0.000225<br>(0.00223) | -0.000324<br>(0.00225) |
| Constant | -3.387<br>(2.201) | -3.384<br>(2.065) | -3.526*<br>(1.969) | -3.156<br>(2.215) | -3.108<br>(2.189) | -3.090<br>(2.141) | -3.427<br>(2.441) |
| Observations | 138 | 138 | 138 | 138 | 138 | 138 | 138 |
| R-squared | 0.401 | 0.406 | 0.407 | 0.407 | 0.407 | 0.412 | 0.412 |
| log Deaths mean | 1.150 | 1.150 | 1.150 | 1.150 | 1.150 | 1.150 | 1.150 |
| log Deaths sd | (1.783) | (1.783) | (1.783) | (1.783) | (1.783) | (1.783) | (1.783) |
| log $BMI \geq 30$ mean | 2.759 | 2.759 | 2.759 | 2.759 | 2.759 | 2.759 | 2.759 |
| log $BMI \geq 30$ sd | (0.623) | (0.623) | (0.623) | (0.623) | (0.623) | (0.623) | (0.623) |

Notes: The dependent variable is log COVID-19 deaths per 100,000 capita. $BMI \geq 30$ and $Age \geq 65$ are the percentage of a population that is severely obese and over 65 years old, respectively. $Days\ cases > 100$ is the number of days since the number of COVID-19 cases is larger than 100. Lockdown response refers to the number of days between the time the number of COVID-19 cases is larger than 100 and the government has introduced a lockdown (i.e., a "containment and health index" $> 0.5$ ). Diabetes, Cardiovascular diseases, and CRDs (chronic respiratory diseases) refer to their prevalence in terms of cases. The GDP per capita is PPP adjusted. Urban population is the percentage of the population living in urban areas. Robust standard errors are in parentheses stars indicating \*\*\* $p < 0.01$ , \*\* $p < 0.05$ , \* $p < 0.1$ .

A country’s economic situation might impact its ability to deal with the COVID-19 pandemic. Hence, GDP per capita is included in our analysis. We control for the share of urban population as in urban areas the SARS-CoV-2 might spread faster due to the higher population density.

Given the dynamic (and lagged) nature of deaths from the pandemic, we purge the estimate from the longer-shorter duration of the epidemic within a specific country controlling for the number of days since the first 100 cases. The idea is simply that of not comparing countries at different horizons of the mortality dynamics. Just to give an example, the first official data is from January 22, 2020, with 548 confirmed cases in China, while in Italy and the US the first 100 cases were passed on February 23 and March 4 respectively. When looking at the development of COVID-19 over time in a single country one can see an increasing death number at an increasing rate at the beginning of the pandemic. As the first wave of COVID-19 fades away, however, the rate decreases. It is, therefore, important to control for the number of days since the first 100 cases as well as its square and cube. Few countries went into a second wave yet (as of August 10, 2020). Looking at the Appendix Figure D.1 we see a robust cubic relationship as expected.

Quick policy responses might be a key factor in keeping the pandemic under control (Hopman et al., 2020). If governments have reacted quickly with lockdown measures after a COVID-19 outbreak this might have been effective at containing the virus. Thus, we control for the number of days between the first 100 cases and a lockdown. For more details on the variables included in the regressions see Appendix Table A.1.

Adding more and more controls to the analysis the results are still there and pretty robust around the 1.5 partial correlation coefficient, importantly when we control for other potential comorbidities the coefficient associated with obesity prevalence varies little and is still significant at the 1% level, while the other ones are much smaller and statistically insignificant. We find the lack of significant correlation between other conditions prevalence and Covid-19 mortality somewhat intriguing as while the different health conditions are highly correlated there still remain a large amount of independent variation left.^3^ Similarly, it appears that the extra controls add little to the analysis, in particular in terms of mortality the share of 65 and older has a small and statistically insignificant effect, while GDP per capita isn’t significantly correlated with mortality (a glimpse of that lack of correlation is also given in Figure 2), a similar consideration applies to the share of urban population.

## 4 Concluding Remarks

We call attention to the relationship between obesity prevalence and COVID-19 mortality rates as very recently also highlighted by the NYT for young men.^4^ While we cannot make any causal statement we note a robust association between obesity prevalence and mortality at the country level for the 138 countries we have reliable data for. We note that it would be crucial at this stage to replicate and expand our analysis with individual-level data, at large scale and across countries, recording BMI and other conditions for all Covid-19 cases. We, therefore, call for systematically collecting and sharing this data. Importantly, and perhaps in line with some recent policy initiatives, we highlight the potential relevance of obesity as a major cofactor associated with heightened mortality risk.^5^ Whether in the short-term focusing on BMI is the relevant strategy to reduce risk of death by or with Covid-19 is beyond the scope of the current paper.

## A Data

**Table A.1:**
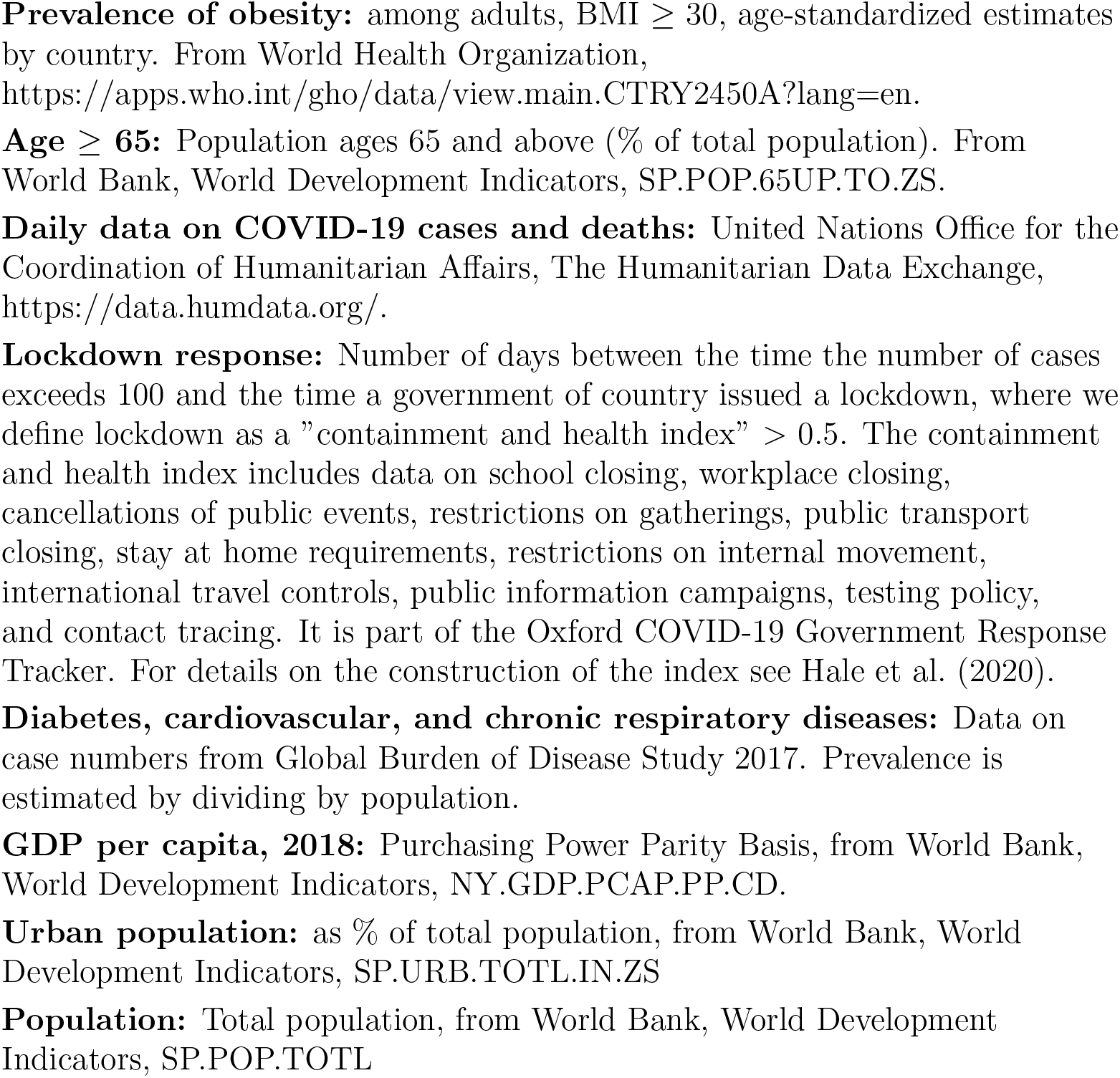
Data

## B. Robustness Checks

### B.1 Dropping 5% Tail Observations on Mortality

**Figure B.1:**
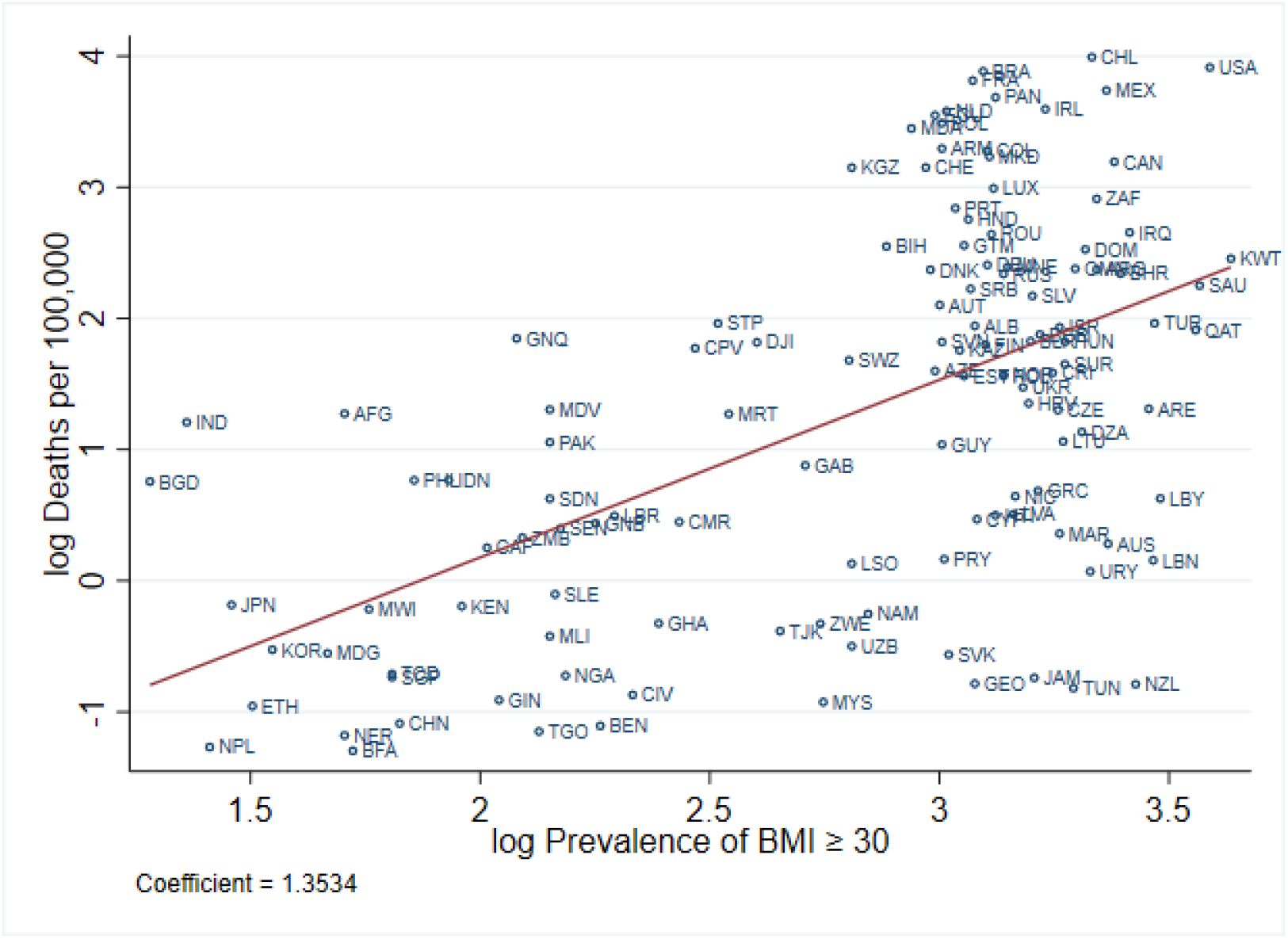
Correlation of Obesity and COVID-19 Deaths. *Notes:* Data on COVID-19 deaths from August 10, 2020. The bottom and top 5% countries with respect to mortality per 100,000 have been dropped.

**Figure B.2:**
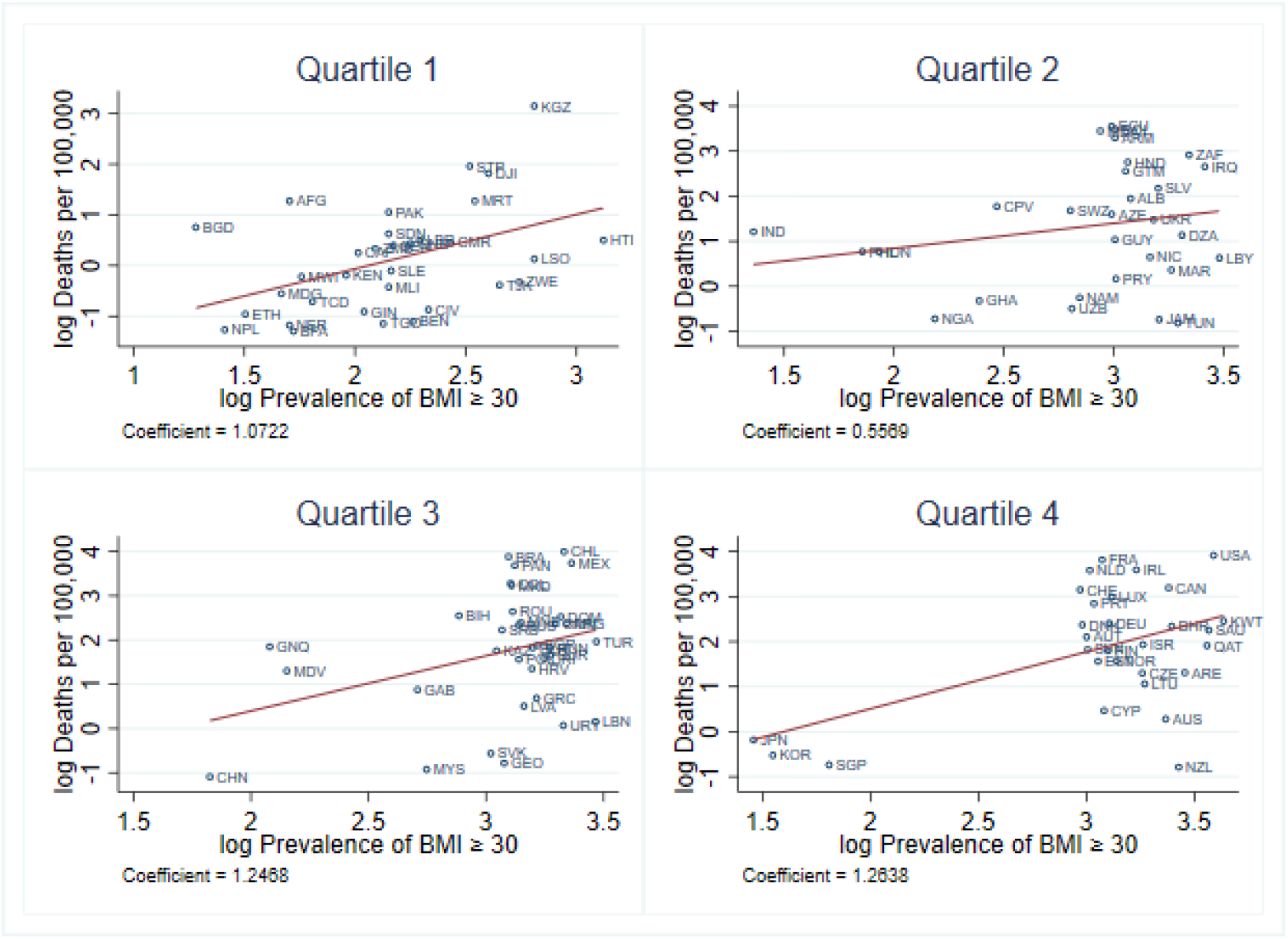
Correlation of Obesity and COVID-19 Deaths. *Notes:* Data on COVID-19 deaths from August 10, 2020. The bottom and top 5% countries with respect to mortality per 100,000 have been dropped.

## C. Europe

**Figure C.1:**
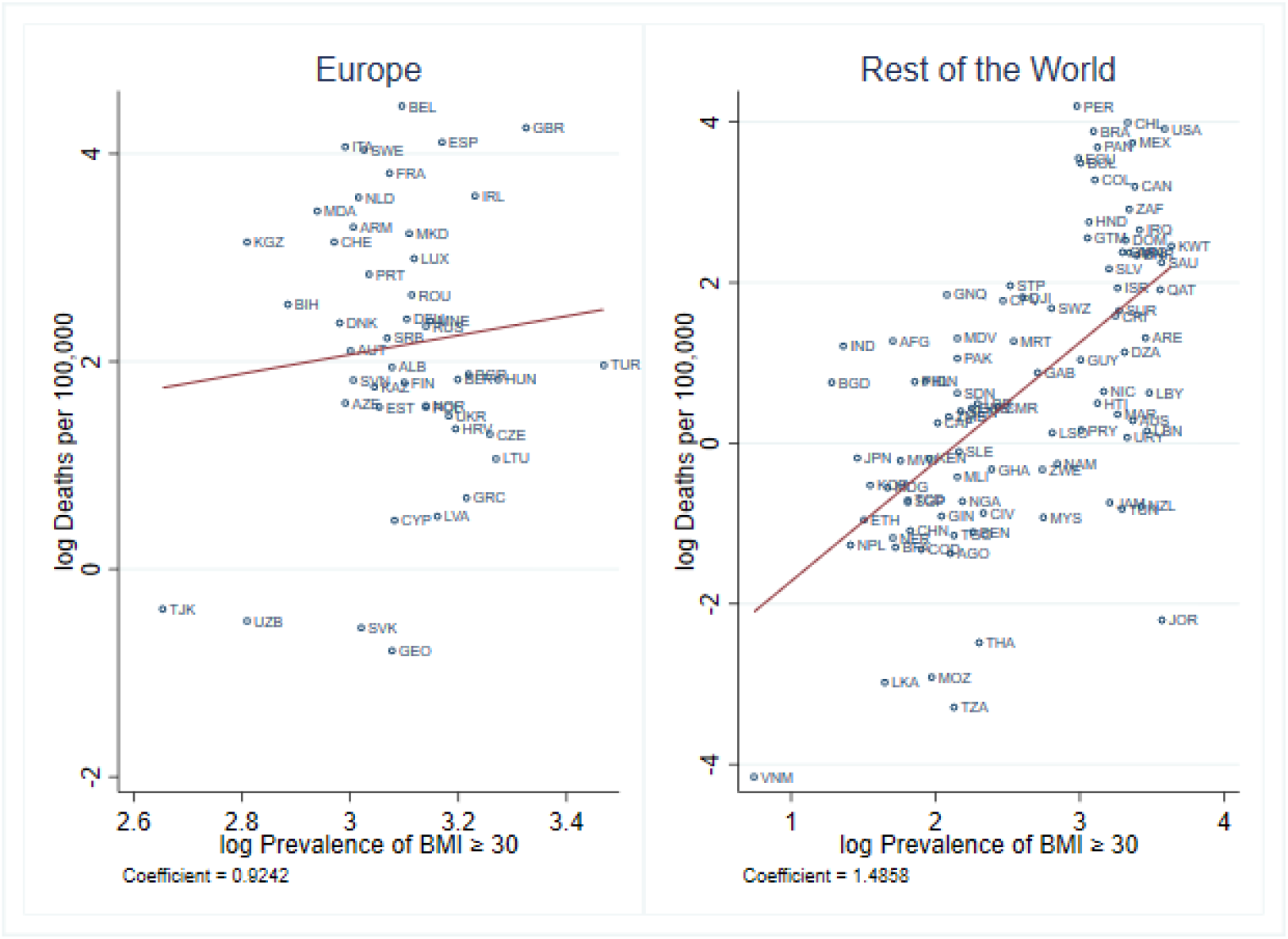
Correlation of Obesity and COVID-19 deaths, Europe vs. The Rest of the World. *Notes:* Data on COVID-19 deaths from August 10, 2020. Europe refers to the countries in “Europe & Central Asia” as defined by the World Bank.

## D. COVID-19 Timeframe

**Figure D.1:**
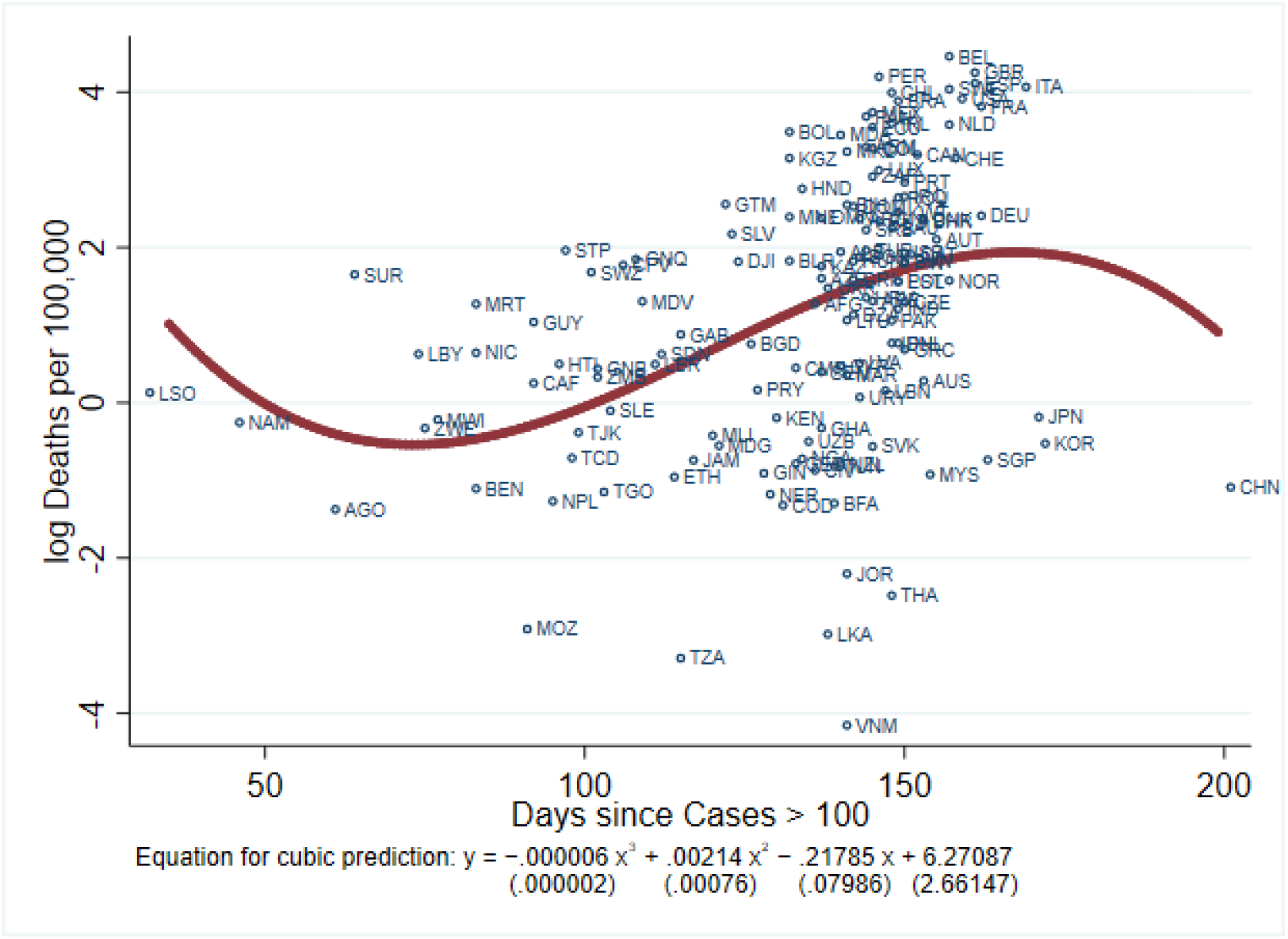
Correlation of COVID-19 Timeframe and COVID-19 Deaths. *Notes:* Data on COVID-19 deaths from August 10, 2020.

## Data Availability

All data publicly available. Please refer to 'Table A.1: Data' for an overview.

## Footnotes

1 https://cepr.org/content/covid-economics-vetted-and-real-time-papers-0

2 Two major studies looking at these patient characteristics are from China Wu and McGoogan (2020) and the Lombardy Region in Italy Grasselli et al. (2020). They do not, however, contain data on the BMI of patients.

3 In a regression of obesity prevalence on all other health conditions and the full set of controls we use in Column (7) of Table 1 the maximum Adjusted R-squared is .67, and importantly in the horse race between different condition obesity prevalence appears to be the leading driver.

4 https://www.nytimes.com/2020/08/14/health/covid-19-obesity.amp.html

5 https://www.theguardian.com/society/2020/jul/11/ no-10-plans-weight-loss-drive-to-ready-uk-for-expected-covid-19-second-wave

## Notes

### Competing Interest Statement

The authors have declared no competing interest.

### Funding Statement

No funding source to be disclosed for this project.

### Author Declarations

No approval needed.

